# Classification of *ACE* variants related to Alzheimer’s disease (AD): the *ACE* mutations — AD browser

**DOI:** 10.64898/2026.08.09.26360046

**Authors:** Anastasiia A. Buianova, Ivan A. Adzhubei, Petr A. Buianov, Olga V. Kryukova, Olga A. Kost, Mikhail Iu. Kuznetsov, Steven M. Dudek, Denis V. Rebrikov, Sergei M. Danilov

## Abstract

**Background:** *ACE* variants are genetic risk factors for Alzheimer’s disease (AD), potentially through reduced enzymatic activity and impaired amyloid-β hydrolysis.

**Objectives:** To create a publicly available database of *ACE* variants relevant to ACE deficiency and AD, and to estimate the population frequency of damaging *ACE* variants and their impact on blood ACE levels.

**Methods:** *ACE* variants were compiled from literature, public databases (VarSome, dbSNP, ClinVar, gnomAD), and sequencing data (WES/WGS) from 5147 Russian individuals. Variants were classified using a consensus *in silico* score (AlphaMissense, MetaRNN, EVE). Blood ACE levels were measured in 330 carriers of 64 different *ACE* mutations.

**Results:** We identified 1682 unique *ACE* variants. Of these, 608 (36.2%) were classified as functionally damaging, including 17 signal peptide, 210 loss-of-function, and 381 missense variants. The estimated carrier frequency of damaging *ACE* variants was 2 % (1/50). Notably, 24 variants associated with experimentally confirmed reductions in blood ACE levels had a combined estimated carrier frequency of 3.9 % in the general population, calculated from cumulative gnomAD v4.1.0 allele frequencies under a rare-variant independence model. An open-access browser is available at https://ace-browser.com/.

**Conclusions:** Variants associated with reduced blood ACE levels were estimated to be carried by approximately 1 in 25 individuals in the general population. This frequency is of the same order of magnitude as the 13.2% prevalence of Alzheimer’s dementia in individuals aged 75–84 years (Alzheimer’s Association, 2025), consistent with the hypothesis that ACE deficiency may represent an underrecognized contributor to late-onset AD susceptibility. The *ACE* mutations-AD browser and integrated genotype-phenotype data presented here provide a novel resource for future basic, translational, and clinical research on ACE-dependent AD.

## INTRODUCTION

Alzheimer’s disease (AD) is the most prevalent neurodegenerative disorder worldwide and constitutes approximately two-thirds of all dementia cases. As of 2019, an estimated 57 million individuals were living with dementia globally, a figure projected to reach 153 million by 2050, driven primarily by population ageing [1]. The economic and societal burden is staggering dementia-associated costs are projected to reach 14.5 trillion INS$ between 2020 and 2050 [2]. Despite decades of intensive research and the recent conditional approvals of amyloid-targeting immunotherapies, no highly effective disease-modifying treatment is broadly available, underscoring an urgent need to identify and characterize novel genetic risk factors which could unlock preventive strategies and precision therapeutic targets.

Large-scale genome-wide association studies have identified several dozen independent risk loci for AD, implicating pathways such as amyloid processing, lipid metabolism, neuroinflammation, endosomal trafficking [3], and many others. However, even these common variants do not fully explain the heritability of late-onset AD (LOAD), and the discovery of rare missense and loss-of-function (LOF) variants within several loci (e.g., *SRC*, *MME*, *TREM2*, *ABCA7*) points toward underexplored risk mechanisms — in particular, rare coding variants in genes which may modulate amyloid-β clearance [4,5].

Among the candidate genes at established GWAS loci, the angiotensin-converting enzyme gene (*ACE*) has attracted sustained attention since the initial reports of its association with AD risk [4,6–9]. *ACE* encodes a zinc metallopeptidase which plays dual physiological roles: it converts angiotensin I to the vasoconstrictor angiotensin II, while it also degrades a diverse repertoire of bioactive peptides, including bradykinin and, critically, the amyloidogenic peptide Aβ42 [10,11]. The enzyme (ACE) is a two-domain ectoprotein (consisting of N- and C-domains), both catalytically active, whose full-length dimeric architecture was only recently resolved at near-atomic resolution by Cryo-EM [12,13].

Crucially, Aβ hydrolysis is preferentially carried out by the N-domain [14], which accommodates larger peptide substrates through an expanded catalytic chamber [15]. This mechanistic specificity positions ACE N-domain activity as a direct, enzymatic brake on amyloid accumulation in the brain. In line with this model, reduced ACE expression or activity is expected to impair Aβ clearance and thereby promote senile plaque formation — a prediction supported by clinical biomarker studies [16,17] — and most recently, by evidence that boosting microglial ACE expression reduces amyloid burden and cognitive decline in the 5xFAD genetically-modified mouse model of AD [18]. Conversely, *ACE* variants which increase ACE catalytic activity or Ang II production may independently promote neuroinflammation and tau hyperphosphorylation [19], illustrating the bidirectional complexity of ACE biology in AD.

Despite this compelling mechanistic framework, the landscape of coding *ACE* variations in the human population has remained incompletely characterized, and its aggregate contribution to AD susceptibility substantially underestimated. We previously analyzed publicly available databases and identified more than 1200 *ACE* variants, of which >400 were predicted to be functionally damaging by consensus *in silico* scoring. Their combined carrier frequency in the general population was estimated at approximately 5%, a figure strikingly comparable to the prevalence of AD in adults older than 70 years [20]. This convergence raised the hypothesis that genetically reduced ACE expression constitutes a common, previously underrecognized risk subgroup for LOAD, analogous in concept to *GBA1* heterozygous variants in Parkinson’s disease [21,22]. Critically, however, the clinical significance of any individual damaging *ACE* variant cannot be inferred from genotype alone. Inter-individual heterogeneity in plasma ACE levels among carriers of the same mutation — likely reflecting the modulatory influence of additional variants at other loci affecting ACE shedding and expression — highlights that comprehensive ACE phenotyping is indispensable for accurate risk stratification [15,23–26].

Here, we present a systematically updated and substantially expanded classification of all catalogued human *ACE* coding variants, integrating data from three complementary sources: curated public databases (VarSome [27], dbSNP, ClinVar, gnomAD v4.1.0), the peer-reviewed literature, and whole-exome and whole-genome sequencing (WES/WGS) data from 5147 individuals of Russian ancestry. We apply a consensus *in silico* pathogenicity framework (AlphaMissense, MetaRNN, EVE) to classify 1682 unique variants — the largest and most systematically annotated *ACE* variant catalogue to date — and estimate the population carrier frequency of damaging variants using gnomAD allele frequency data. We further report measured plasma ACE levels in 330 carriers of 70 distinct *ACE* mutations, revealing that approximately 1 in 14 individuals in the general population carries a variant associated with reduced blood ACE. This prevalence is of the same order of magnitude as AD dementia in individuals aged 75–84 years [28]. Using a broad panel of conformation-specific monoclonal antibodies (mAbs), we identify antibody-based markers for 19 distinct *ACE* mutations and demonstrate that conformational fingerprinting can detect ACE-deficient variants even in the absence of sequencing data — an approach of particular value in resource-limited settings and for future therapeutic monitoring. To facilitate open science, all variant-level data, genotype–phenotype associations, and structural annotations are made freely accessible through a dedicated online browser. Together, our findings delineate a large and underappreciated genetic risk stratum for AD, provide a mechanistic framework for ACE-dependent AD susceptibility, and lay the groundwork for genotype-informed prevention strategies and targeted pharmacological rescue of traffic-deficient *ACE* variants.

## METHODS

### Literature Search Criteria

We searched for all reported coding variants of the human *ACE* gene (gene ID: 1636; reference transcript NM_000789.4) using three complementary approaches.

First, we queried the VarSome clinical genomics platform (https://varsome.com) [27] using its Region Browser tool. We focused on variants which had at least one submission from curated sources, including ClinVar, UniProt, and User-Linked articles (peer-reviewed publications). This ensured that only variants with documented evidence were included.

Second, to ensure completeness, we performed an independent search in dbSNP using the following query: «ACE[All Fields] AND coding_sequence_variant[Function Class] NOT non_coding_transcript_variant[Function Class]». This dbSNP search yielded 1504 unique entries. It was narrowed to stop-gained variants with the query: «ACE[All Fields] AND stop_gained[Function Class] NOT non_coding_transcript_variant[Function Class]», which identified 77 unique stop-gained variants.

Third, we manually searched the peer-reviewed literature (PubMed, Google Scholar) for original reports describing *ACE* variants, including case studies, association studies, and functional analyses. Whenever a variant was reported in the literature, we recorded the corresponding reference number. All variants from the three sources were filtered to retain only those located in the coding region (exons 1–26) and canonical splice sites (±2 bp) of NM_000789.4 (except c.1709+5G>T). Variants reported exclusively in non-canonical transcripts or lacking unambiguous genomic coordinates were excluded. Where discrepancies between VarSome and dbSNP were found (e.g., redundant entries, variants annotated to different transcripts, or multinucleotide variants), all records were manually reconciled. The final list contained 1682 unique *ACE* variants.

### Identification of ACE mutations in the available cohorts of sequenced subjects

To supplement literature-derived variants, we analyzed whole-exome sequencing (WES) data from 5147 individuals of Russian ancestry. These samples originated from two cohorts:

(1) Pirogov Russian National Research Medical University (RSMU) cohort — 4657 individuals. DNA libraries were prepared using the MGIEasy Universal DNA Library Prep Set (MGI, Shenzhen, China) and enriched with SureSelect Human All Exon v7 baits (Agilent Technologies, Santa Clara, CA, USA) according to the “RSMU exome” protocol as previously described [29]. Sequencing was performed on DNBSEQ G-400 (MGI Tech) in PE100 mode to target mean coverage >100×. Data processing used an automated Python3 pipeline: quality assessment with FastQC v0.12.1 [30], read trimming with BBDuk v38.96 [31], alignment to GRCh38/hg38 using bwa-mem2 v2.2.1 [32], SAM-to-BAM conversion and sorting with Samtools v1.9 [33], duplicate marking with Picard v2.22.4 [34], variant calling with BCFtools v1.9 [35] and DeepVariant v1.5.0 [36], normalization with vt normalize v0.5772 [37], and annotation with InterVar v2.2.2 [38].

(2) Institute on Aging Research (Russian Gerontology Research and Clinical Center) cohort — 490 participants. Whole-genome sequencing and initial processing were performed as previously described [25]. Fastq files were processed with FastQC v0.11.9 [30], trimmed with BBDuk v38.96 [31], aligned to GRCh38 using bwa-mem2 v2.2.1 [32], and duplicates removed with Picard MarkDuplicates v2.22.4 [34]. VCF files were obtained using VarAFT v2.17 [39] with dbSNPv150 and annotated with rsIDs from dbSNPv156 using bcftools v1.18 [35].

The *ACE* I/D polymorphism was identified based on the linked rs4343. All identified variants from both cohorts were merged, normalized, and cross-referenced with VarSome and gnomAD v4.1.0. Multinucleotide variants which could not be reliably phased were excluded from further analysis.

### Classification of ACE mutations

After compiling the complete list of 1682 unique *ACE* variants from the literature, VarSome, dbSNP, and our sequencing cohorts, we performed a uniform *in silico* annotation to assess their potential functional impact. To this end, we created a pseudo-VCF file containing all identified variants and submitted it to the Variant Effect Predictor (VEP, Ensembl release 113) [40]. VEP was used to retrieve AlphaMissense and MetaRNN scores for all variants, EVE scores for missense variants only, and the predicted nonsense-mediated decay (NMD) escape status for frameshift and nonsense variants. For variants located in the signal peptide-encoding region (87 variants, Table S1), pathogenicity was assessed using AlphaMissense and MetaRNN. Each score was converted to a rank (pathogenic = 3, ambiguous = 2, benign = 1), and the arithmetic mean of the two ranks was calculated. A mean greater than 2.5 was defined as pathogenic, a mean greater than 1.5 but less than 2.5 as ambiguous (VUS), and a mean of 1 as benign.

For missense variants in the mature protein (1,385 variants, Table S1), all three scores (AlphaMissense, MetaRNN, and EVE) were used. Each was converted to the same 3-point rank scale (pathogenic = 3, ambiguous = 2, benign = 1), and the mean of the three ranks was calculated. A mean of 2.5 or higher was defined as pathogenic, a mean greater than 1.3 but less than 2.5 as ambiguous (VUS), and a mean of 1.3 or lower as benign.

Frameshift, indels and nonsense variants (210 variants, Table S1) were considered LOF. In addition, their predicted escape from nonsense-mediated decay was obtained from VEP (highlighted in Table S1). Splice site variants (Table S2) were analyzed separately with SpliceAI [41], SAI-10k-calc [42], and SpliceVault [43] to predict the most likely splicing outcome.

All consensus classifications, together with gnomAD v4.1.0 allele frequencies and measured blood ACE levels (where available), are reported in Table S1 and summarized in the Results.

### Estimation of carrier frequency for ACE variants

For each category of variants, we estimated the proportion of individuals carrying at least one damaging variant (Gene Carrier Rate, GCR). Assuming that damaging variants are rare (MAF < 0.001) and occur independently, the probability of carrying a specific variant can be approximated as *2MAF_i_*. Therefore, the probability of not carrying any damaging variant within a given category is 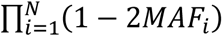, where *N* is the number of damaging variants in the category. The corresponding GCR is 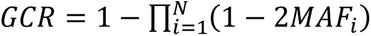.

Here, *MAF_i_* denotes the minor allele frequency of variant *i*, obtained from gnomAD v4.1.0. This approximation is appropriate because all included variants are rare, no homozygous carriers were reported in gnomAD, and strong linkage disequilibrium (LD) between variants was not observed. The probability of compound heterozygosity is therefore expected to be negligible.

### Blood ACE phenotyping (estimation and measurement)

Blood ACE phenotyping was performed on plasma (primarily EDTA-plasma but occasionally sera) samples from 330 carriers of 62 ACE mutations and 500+ corresponding controls. Blood samples were obtained from previously sequenced (WES/WGS) subjects from two primary sources: the Center for Precision Genome Editing and Genetic Technologies for Biomedicine (*n* = 4657) and the Institute on Aging Research, Russian Gerontology Research and Clinical Center (*n* = 490). Both belong to the Pirogov Russian National Research Medical University, Moscow, Russia, and were taken from previously published studies [23–26]. The collection of human blood samples and the protocol for the whole exome screening of tested individuals were reviewed and approved by the Ethics Committee. All subjects gave written informed consent for the use of any data for scientific purposes. Measured blood ACE values from subjects with the corresponding ACE mutations were measured as median values.

Blood ACE levels were measured in plasma (and occasionally in available sera) samples using precipitation of ACE protein by mAb 9B9 followed by measurement of precipitated ACE activity directly on the bottom of the 96-well plates using artificial short substrates (ZPHL and HHL) as previously described [23,44,45].

In some experiments, ACE protein was also precipitated from plasma samples using other mAbs from our collection of more than 40 different mAbs to different epitopes of ACE, and mAb binding ratios were then calculated (i.e., conformational fingerprinting of ACE) [46–48]. This approach allowed us to identify some combinations of mAbs as specific markers for 19 different mutations of *ACE*.

### Construction of browser and data sharing

We developed an open-access, web-based interactive *ACE* variant browser using a Python v3.12-based backend implemented with FastAPI v0.115.0 and an asynchronous SQLite v0.20.0 database layer managed through SQLAlchemy v2.0.36. The client interface was implemented as a static HTML, CSS and JavaScript application, with Tailwind CSS v3.4.17 used for stylesheet generation, Leaflet v1.9.4 for map visualization, and NGL.js v2.0.0-dev.39 used for interactive protein-structure visualization. The browser was designed to support searchable and filterable exploration of curated *ACE* variant data through dedicated API endpoints.

For each variant, the platform integrates multilayered annotations, including HGVS-compliant genomic and protein-level nomenclature, GRCh38-mapped genomic coordinates, amino acid positions referenced to the canonical ACE isoform, protein domain assignments, rsID identifiers where available, and population allele-frequency information derived from gnomAD v4.1.0. Variant-level annotations also include *in silico* pathogenicity metrics, including CADD, MetaRNN, EVE and AlphaMissense scores, together with ClinVar clinical classifications where available.

Each variant was assigned a functional category reflecting its presumed biological consequence, including catalytic, shedding-associated, dimerization-associated or structural effects. Where available, variant records were linked to blood ACE phenotype data obtained from published cohorts or institutional measurements, together with corresponding primary bibliographic sources. The underlying curated dataset is stored in structured tabular form and exposed through the browser API to enable reproducible retrieval, filtering and downstream reuse. The resource was designed for iterative updating, allowing newly reported *ACE* variants, revised annotations, gnomAD-derived population-frequency data and additional phenotype information to be incorporated as they become available.

## RESULTS and DISCUSSION

### 1. Identification and classification of *ACE* variants

We identified 1682 unique *ACE* variants (Table S1). The variants were stratified into three functional categories: signal peptide region (residues 1–29 of pre-ACE), frameshift/nonsense, and missense. Within the signal peptide-encoding region, we found 87 unique genomic variants: 61 missense, 6 frameshift, 1 nonsense, and 19 in-frame indels. At the protein level, after accounting for synonymous codons, the 61 genomic missense variants collapsed to 58 distinct amino acid substitutions. Using the consensus pathogenicity scores (AlphaMissense + MetaRNN), we identified 17 damaging variants (10 missense, 6 frameshift, 1 nonsense) with a combined allele frequency of 640 per 100,000 (0.0064) in gnomAD v4.1.0. Their estimated GCR was 1.3 % (approx. 1 in 79 individuals). The most common damaging missense was p.Met1Arg (rs1005792910, 0.8 per 100,000). The remaining 70 variants were benign or ambiguous (VUS).

Integration of splice-site variant annotation (Table S2) revealed a separate class of high-impact regulatory variants. These are characterized by exon skipping, cryptic splice site activation, or intron retention, frequently resulting in predicted loss-of-function via nonsense-mediated decay or in-frame structural remodeling of ACE isoforms.

In the mature ACE coding sequence, we catalogued 210 unique genomic LOF variants (frameshifts, in-frame indels, and nonsense). After collapsing genomic events which lead to identical protein-level changes, the number of distinct protein alterations was 205. Their combined allele frequency was 82 per 100,000 (0.000825), yielding a GCR of 0.17 % (approx. 1 in 607). The most frequent was p.Gly1174AlafsX12 (rs754265941) at 21 per 100,000. Nine of these 210 variants were predicted to escape NMD.

At the nucleotide level, we detected 1,385 unique missense variants, which represented 1,358 distinct missense changes in the ACE protein after collapsing by the encoded amino acid (Table S1). Using a consensus score (AlphaMissense, MetaRNN, EVE; mean rank ≥2.5 = pathogenic), we classified 381 nucleotide variants (374 unique amino acid changes) as pathogenic. Their combined allele frequency was 271 per 100,000 (0.00271), corresponding to a GCR of 0.54% (approx. 1 in 185). The remaining 1,003 nucleotide variants were benign (607) or VUS (396). The most frequent pathogenic missense was p.Pro351Leu (rs2229839) at 35 per 100,000.

Combining all damaging variants from the three categories (17 signal peptide, 210 frameshift/nonsense/indel, and 381 pathogenic missense), the total allele frequency was 994 per 100,000 (0.00994). Assuming rare and independent variants (no strong LD and negligible contribution of homozygous carriers), the proportion of individuals carrying at least one damaging *ACE* variant was estimated using the Poisson limit of the exact product formulation:

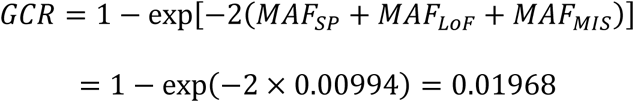

To facilitate practical variant interpretation, the 50 most frequent *ACE* coding variants are summarized in Table S3, ranked by population allele frequency and annotated with functional classification and available plasma ACE phenotype data. This resource enables clinicians to quickly evaluate whether an observed *ACE* variant represents a common population allele and whether previous studies have linked it to altered ACE activity or AD risk.

Thus, approximately 1 in 50 individuals carries a potentially damaging *ACE* variant. This prevalence is similar to the lifetime risk of LOAD in individuals over 75 years (5–10%), consistent with the hypothesis that ACE deficiency is a common, underappreciated genetic risk factor that likely requires additional environmental or genetic hits for disease manifestation.

### 2. Characterization of *ACE* mutations

#### Catalytic architecture of ACE

Cloning of the human *ACE* gene revealed the presence of two conserved HExxH motifs, suggesting that there might be two catalytic centers [49]. Both the N- and C-domains were subsequently shown to contain functional catalytic sites [50], a finding later confirmed by X-ray crystallography of isolated domains [51,52].

Extensive structural and mutagenesis studies, particularly from the groups of Acharya and Sturrock and supported by more than 60 ACE-ligand complex structures, have substantially expanded the catalogue of residues contributing to ACE function [53]. Note that these ACE-ligand combinations include N-domain complexes with amyloid-β42 fragments, linking ACE-mediated peptide hydrolysis to AD–relevant pathways [54].

Beyond residues directly involved in catalysis [55] and the coordination of chloride as an allosteric activator [56], structural and mutagenesis studies have identified the determinants of substrate specificity and domain-selective activity [53, 57–59]. While the active centers on the two ACE domains are quite similar, they play overlapping albeit distinct roles. Thus, the C domain predominantly hydrolyzes angiotensin I and plays a crucial role in the regulation of blood pressure, while the N domain serves as a primary site for the hydrolysis of AcSDKP, which has been implicated as anti-fibrotic agent [57,60–62]. This highlights the need for the development of domain-specific inhibitors. С domain-specific inhibitors would reduce adverse effects related to elevated levels of substance P and bradykinin, while retain an ability of ACE to hydrolyse Aβ42, and to slow Alzheimer’s disease development. N domain-specific inhibitors could reduce inflammation and increase anti-fibrotic capacity by not cleavage of AcSDKP. The potential contribution of ACE deficiency to the development of AD [20] raises concerns about using ACE inhibitors in elderly patients to treat hypertension, especially for those with AD, or as preventive therapy for AD [63]. Recent cryo-electron microscopy structures of full-length glycosylated somatic ACE have further clarified domain organization and inter-domain structural coupling [12,13].

Integration of structural and functional datasets has identified 49 residues in the N-domain and 29 residues in the C-domain whose substitutions are predicted to affect catalytic performance (Table S4), corresponding to a total of 78 residues with functional relevance to enzymatic activity. The N-domain contains a higher number of residues in this analysis, consistent with its previously described involvement in amyloid-β42 hydrolysis [14,15].

#### Structural determinants of ACE dimerization

ACE dimerization occurs primarily through interactions involving the N-domain. Initial evidence was obtained in 2000 using a reverse micelle membrane model in which somatic ACE was shown to form dimers [64]. ACE dimerization was completely inhibited by Neu5Ac- or Gal-terminated saccharides, with the most effective being 3’SiaLac, especially the whole pool of ACE oligosaccharide chains [64]. These highly specific interactions demonstrated the existence of a carbohydrate-recognizing domain on the surface of ACE globule. The exact amino acid residues forming the carbohydrate-recognizing domain on the N domain have not yet been determined. However, fine epitope mapping of ACE defined the general region on the N domain globule, which was shielded by two partially competitive antibodies, 9B9 and 3G8, preventing ACE dimerization [65]. The region may involve amino acid residue Asn82 bearing an oligosaccharide chain, which could participate in dimerization [65,66]. This hypothesis was later confirmed by cryo-electron microscopy structures of full-length glycosylated somatic ACE [12,13]. These studies provided not only structural evidence that somatic ACE dimerization occurs via N domain interface in a native state, but they also revealed a glycan-mediated bridge between N82-linked glycans. Glycan-glycan interactions are known to be very weak and usually require whole clusters of glycan moieties that precede tighter protein-protein interactions [67]. Therefore, cryo-EM is likely to succeed in revealing the first step that occurs in ACE-ACE interactions.

In addition to this glycan–glycan interface, structural analyses identified a central role for Y465 (corresponding to the Y465D mutant in a multigenerational Wichita cohort), in which carriers exhibited a ∼4-fold increase in circulating ACE levels, likely reflecting altered shedding dynamics [68]. Extended structural interrogation of 23 X-ray structures of truncated N-domain dimers further suggested a broader dimerization interface comprising 23 residues [69], involving close packing of the C-loop-2 and C-loop-3 regions of opposing protomers [12]. More recently, high-resolution cryo-EM structures of full-length ACE dimers have indicated that inter-monomer interactions are additionally stabilized by contacts between D2 subdomains of one protomer and D4 subdomains of the opposing protomer [13].

Integration of structural datasets and docking simulations led to the identification of two partially overlapping sets of residues potentially involved in ACE dimerization: 61 residues in the N-domain derived from structural analyses (Table S5a), and 47 residues derived from docking-based predictions (Table S5b), including three residues in the C-domain also implicated in dimer formation [13]. Variants within these interfaces may modulate the extent of ACE dimerization, thereby influencing circulating ACE levels through altered shedding efficiency [69]. In addition, they may affect inter-domain orientation and fine catalytic properties, including inhibitor binding affinity and substrate specificity, particularly for large peptide substrates such as amyloid-β42 [12].

#### ACE mutations that may influence ACE shedding

Both *ACE* genotype and phenotype may contribute to AD–related risk assessment, as circulating ACE levels represent an integrated outcome of genetic variation and post-translational processing [26]. Accordingly, variants which substantially alter ACE shedding have potential clinical relevance, as they directly influence measured plasma ACE concentrations. We identified 38 *ACE* variants associated with altered shedding (Table S6). These variants are predominantly located within the stalk region and are therefore positioned to affect cleavage by the putative ACE secretase at the juxtamembrane interface. Alterations in this region may reduce or enhance proteolytic ectodomain release and, in extreme cases, disrupt membrane anchoring, resulting in constitutive secretion of ACE into the circulation [69].

### 3. Blood ACE phenotyping in available blood samples

Blood ACE activity was assessed across multiple variant carriers using datasets compiled in Tables S1, S7, and S8. Table S1 summarizes 1682 *ACE* variants and includes available plasma ACE measurements for carriers of 74 variants, alongside estimated values for an additional 45 variants. Table S7 reports directly measured plasma ACE concentrations in 330 individuals from the RSMU cohort, representing carriers of 62 distinct *ACE* variants.

Across these datasets, 72 variants were associated with significantly reduced plasma ACE levels (Table S8). Notably, the combined allele frequency of variants associated with decreased circulating ACE was approximately **3.9** % in the studied population. This indicates that genetically determined reductions in ACE abundance are not rare and may represent a substantial fraction of population-level variability in ACE phenotypes. Among recurrent *ACE* variants, Y215C, Q259R, G325R, T352M, I759V, T887M, and N1007K were consistently associated with reduced plasma ACE levels across the analyzed cohorts (Tables S1, S7, and S8). In several cases, reduced circulating ACE is most plausibly explained by impaired protein maturation and reduced cell-surface expression, consistent with a transport-deficiency mechanism previously described for ACE Q1069R [70]. This paradigm— whereby altered processing and trafficking of amyloid-degrading enzymes contributes to impaired Aβ clearance—is increasingly recognized as a critical component of the amyloid clearance deficit in late-onset Alzheimer’s disease, as comprehensively reviewed by Nalivaeva and Turner [9].

Variant Y215C was observed in 84 carriers [26] and demonstrates a pronounced decoupling between genotype and phenotype. Although carriers of this variant as a group exhibited reduced plasma ACE levels (median 62.4% of the mean value of blood ACE in a healthy population, taken as reference 100%), inter-individual variability was substantial. Comparable heterogeneity was observed for other recurrent variants with multiple carriers [15,23,25]. These findings indicate that plasma ACE concentration is not fully determined by genotype alone but reflects additional factors including protein folding efficiency, intracellular trafficking, and post-translational processing.

Collectively, these observations support a systems-level model in which *ACE* variants exert their phenotypic effects primarily through perturbations of protein maturation and cell-surface expression rather than isolated effects on catalytic function. Consequently, genotype-based interpretation alone may be insufficient for accurate functional classification of ACE deficiency states. Integrated assessment is required that combines sequencing-based variant identification with quantitative plasma ACE phenotyping to avoid misclassification of functional status. Across the combined datasets, variants associated with reduced plasma ACE levels account for approximately **3.8**% of the studied population (Table S8), indicating that genetically and functionally relevant modulation of ACE abundance represents a relatively common phenotype.

#### Epitope-resolved characterization of mutant ACE

To further resolve functional heterogeneity among ACE variants, we applied a panel of more than 40 mAbs recognizing conformational epitopes distributed across the N- and C-domains of ACE [47,48,71]. This antibody set has previously enabled detection and structural–functional characterization of multiple *ACE* variants in recombinant systems and in human carriers [46,68,72,73]. Using epitope mapping of mutation-containing regions, we identified antibody-binding signatures defining 19 *ACE* variants (Table S9; Fig. 1). These signatures reflect mutation-induced conformational perturbations, including alterations consistent with impaired folding and reduced cell-surface expression. Importantly, antibody-based detection provides a functional layer of information which is partially independent of sequence-based annotation, enabling phenotypic stratification of *ACE* variants at the protein level.

**Fig. 1.**
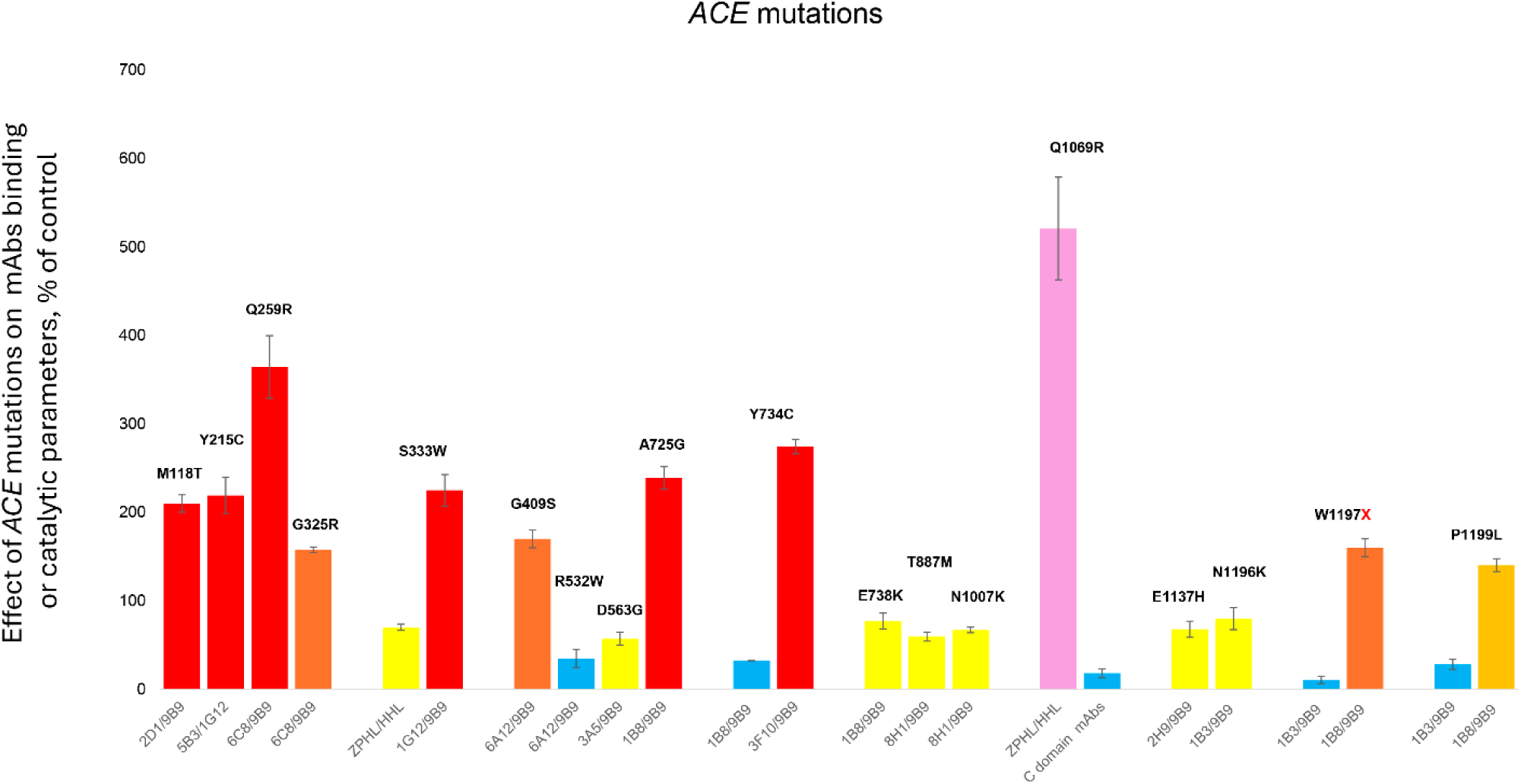
Conformational fingerprinting of 19 *ACE* variants by mAb binding ratios and catalytic assays. Each bar represents the effect of the indicated *ACE* mutation on the binding ratio of a specific mAb pair (e.g., 6C8/9B9) or on the ZPHL/HHL catalytic ratio, expressed as percentage of the mean value in healthy controls (100%). Bar colors reflect the magnitude of deviation from control: red, >200%; orange, 150–200%; brown, 120–150%; gray, 80–120%; yellow, 50–80%; blue, <50% of control values. The lilac-colored bar (Q1069R, ZPHL/HHL) reflects an elevated N-domain-to-C-domain catalytic activity ratio rather than an antibody binding parameter and is shown separately to indicate a qualitatively distinct readout. Error bars represent standard deviations. Data are derived from Table S9.

From a translational perspective, this approach may support identification and monitoring of transport-deficient *ACE* variants in settings where sequencing is limited, or where protein-level readouts are required. In principle, such antibody-defined signatures may also enable longitudinal monitoring of therapeutic strategies aimed at restoring proper folding and trafficking, as previously demonstrated for the transport-deficient Q1069R variant *in vitro* [70].

### 4. *ACE* mutations and AD

Beyond its canonical role in blood pressure and vascular tone regulation [74,75], and its emerging functions in immune regulation [76], ACE has increasingly been implicated in the molecular pathology of AD [4,6,7,18,20, 77]. To systematically assess the contribution of *ACE* genetic variation to AD-related biology, we extracted a curated subset of 22 *ACE* variants previously reported to be associated with AD from the full variant compendium (Table 1). Importantly, these variants are distributed across multiple functional classes, including residues implicated in catalytic regulation, protein folding, trafficking, and extracellular processing, suggesting mechanistic heterogeneity rather than a single unified pathogenic pathway.

**Table 1.** *ACE* variants previously reported in association with Alzheimer’s disease and their population frequency in the study dataset. Notes: The p.Ser1238ProfsX42 variant escapes nonsense-mediated decay (NMD). MAF color coding: Values >10 per 100,000 alleles are shown in bold; ≥100 in red; ≥1000 in bold red. Abbreviations: MAF – minor allele frequency (gnomAD v4.1.0); AD – Alzheimer’s disease; VUS – variant of uncertain significance; LOF – loss-of-function. Blood ACE levels are color-coded by deviation from control (100%): red >200%; orange 150–200%; brown 120–150%; gray 80–120%; yellow 50–80%; blue <50%. References in brackets/square parentheses denote original AD association reports.

| HGVSp | Amino acid residue | rsID (reference) | MAF /100 000 gnomAD v.4.1.0 | Consensus pathogenicity | Blood ACE, % of control | AD |
| --- | --- | --- | --- | --- | --- | --- |
| Frameshifts and nonsense |  |  |  |  |  |  |
| p.Arg149LeufsX54 | R120LfsX54 | rs778759606; [17,78] | 9.2 | Pathogenic (LOF) | Low | AD [17] |
| p.Trp343X | W314X | rs200225958; [17,78] | 0.4 |  | Low |  |
| p.Arg442ValfsX14 | R413VfsX14 | rs1442562714; [17] | 0.1 |  | - |  |
| p.Ser1027TyrfX14 | S998YfsX14 | rs1170915014; [17] | 0.1 |  | - |  |
| p.Asp1058TyrfX16 | D1029YfsX16 | [17] | 0 |  | Low |  |
| p.Ser1238ProfsX42 | S1209PfsX42 | [17] | 0 |  | Low |  |
| Missense |  |  |  |  |  |  |
| p.Asn160Ser | N131S | rs117134739; [79] | 49 | Benign | 74 | AD [79] |
| p.Tyr244Cys | Y215C | rs3730025; [4,7,80] | 1394 |  | 62 [26] | AD [4, 7] |
| p.Gly354Arg | G325R | rs56394458; [7] | 853 |  | 78 | AD [7] |
| p.Thr381Met | T352M | rs150466411; [7] | 104 |  | 73 | AD [7] |
| p.Arg561Trp | R532W | rs4314; [7,72,81] | 57 |  | 500 [72] | AD [7] |
| p.Asp592Gly | D563G | rs12709426; [7,82,83] | 279 |  | 106 [84] | AD [7,83] |
| p.His629Pro | H600P | rs201594771; [7] | 54 |  | 160 | AD [7] |
| p.Ser660Cys | S631C | rs147429960; [7] | 144 |  | 111 [24] | AD [7] |
| p.Glu767Lys | E738K | rs148995315; [7] | 36 | VUS | 131 [15] | AD [7] |
| p.Ile798Val | I769V | rs117647476; [7] | 350 | Benign | 79 | AD [7] |
| p.Thr916Met | T887M | rs3730043; [7,83] | 655 |  | 79 | AD [7,83,85] |
| p.Gly1013Ser | G984S | rs571848794; [7,86] | 6.1 | Pathogenic | - | AD [7] |
| p.Ile1018Thr | I989T | rs4976; [7,70,87] | 34 | VUS | 188 [24] | AD [7] |
| p.Asn1036Lys | N1007K | rs142947404; [7,83] | 64 | Benign | 58 [25] | AD [7,83,85] |
| p.Ala1252Val | A1223V | rs762056936; [88] | 0.7 |  | - | AD [88] |
| p.Arg1279Gln | R1250Q | rs4980; [7,83] | 527 |  | 86 | AD [7,83] (♀) |
| Sum: |  |  | 4547 |  |  |  |

Notes: The p.Ser1238ProfsX42 variant escapes nonsense-mediated decay (NMD). MAF color coding: Values >10 per 100,000 alleles are shown in bold; $\geq 100$ in red; $\geq 1000$ in bold red. Abbreviations: MAF – minor allele frequency (gnomAD v4.1.0); AD – Alzheimer's disease; VUS – variant of uncertain significance; LOF – loss-of-function. Blood ACE levels are color-coded by deviation from control (100%): red >200%; orange 150–200%; brown 120–150%; gray 80–120%; yellow 50–80%; blue <50%. References in brackets/square parentheses denote original AD association reports.

Based on aggregated allele frequency estimates derived from Table 1, the combined population prevalence of AD-associated *ACE* variants exceeds 4.5%. This indicates that a substantial fraction of the general population carries at least one *ACE* variant previously linked to AD-relevant molecular mechanisms. From a clinical genetics’ perspective, this observation is critical. The presence of an *ACE* variant annotated as AD-associated does not imply deterministic disease risk but rather contributes to a polygenic and context-dependent landscape in which ACE function is modulated at multiple biological levels, including enzymatic activity, membrane expression, and circulating protein abundance. Consequently, interpretation of *ACE* variation in AD must integrate both genotypic annotation and phenotypic readouts of ACE function.

These findings support a model in which ACE-related AD susceptibility arises from distributed perturbations across the ACE functional network, rather than from isolated high-penetrance mutations. This reinforces the importance of integrating population-scale variant burden with functional ACE phenotyping when evaluating the potential contribution of ACE to AD risk.

### 5. *ACE* variant browser implementation and web interface

The browser integrates five analytical layers: (i) sequence-level 1682 *ACE* variant distribution (Table S1), (ii) functional *ACE* mutations annotation (Tables S4–S6), (iii) population frequency mapping (gnomAD v4.1.0), (iv) plasma ACE phenotyping (Table S7, S8), and (v) mAbs-based conformational profiling (Table S9) (Fig.2).

**Fig 2.**
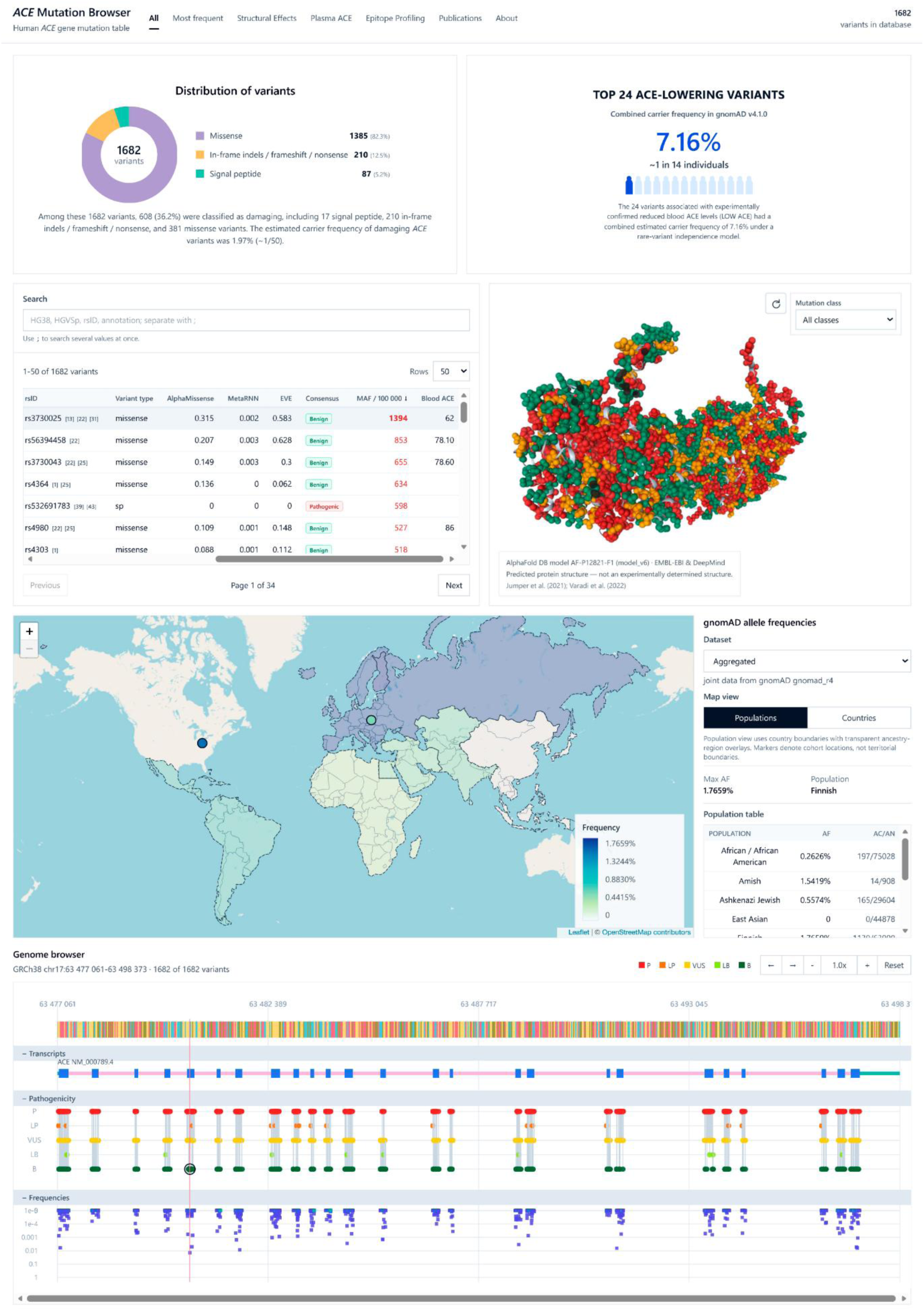
Overview of the *ACE* mutations–AD browser. The browser integrates *ACE* variant annotation, population frequency data (gnomAD v.4.1.0), protein structure visualization, plasma ACE phenotyping, and antibody-based conformational profiling into a unified interactive platform. Available at https://ace-browser.com/

Global mapping of variants along the *ACE* coding sequence revealed a non-uniform distribution of genetic variation across the protein. Distinct regional enrichment patterns were observed corresponding to major functional modules, including catalytic domains, dimerization interfaces, membrane-proximal shedding regions, and splice-sensitive loci. These patterns indicate that *ACE* variation is structurally and functionally compartmentalized rather than randomly distributed across the gene.

Stratification of variants by predicted functional consequences demonstrated concordant clustering of catalytic-impact variants within both N- and C-domain active-site regions (Table S4), consistent with known enzymatic architecture. Variants assigned to dimerization-associated classes (Tables S5a–S5b) were preferentially localized to N-domain interface regions, including residues implicated in glycan-mediated contacts and inter-protomer packing surfaces. Shedding-associated variants (Table S6) were predominantly confined to the stalk and transmembrane-proximal regions, consistent with their proposed role in regulating ectodomain release.

Overlay of population frequency data from gnomAD v4.1.0 demonstrated that functionally relevant *ACE* variation is not restricted to rare alleles. Instead, recurrent variants contribute measurably to the overall allelic architecture of *ACE*, including variants previously associated with AD (Table 1). Collectively, these variants represent a non-trivial proportion of population-level *ACE* genetic diversity (>4.5% in aggregate), indicating that functional variation in *ACE* is widespread at the population scale.

Integration of genotype with available phenotypic data (Tables S7–S8) further revealed substantial heterogeneity in circulating ACE levels among carriers of recurrent variants. While several variants were consistently associated with reduced plasma ACE, inter-individual variability was observed within each genotype class, indicating that genotype alone does not fully determine systemic ACE abundance. Finally, integration of antibody-based epitope profiling (Table S9) enabled functional resolution of conformational effects for a subset of variants, linking sequence variation to altered epitope accessibility and supporting a mechanistic connection between structural perturbation and protein-level phenotypes. Collectively, browser-based analysis revealed that *ACE* genetic variation is organized into partially overlapping functional layers governing enzymatic activity, protein trafficking, membrane processing, and conformational stability, rather than existing as isolated variant effects.

## CONCLUSIONS

1. We identified 1682 unique *ACE* variants and classified 608 of them as potentially damaging, establishing the most comprehensive *ACE* variant resource currently available.
2. Damaging *ACE* variants (predicted *in silico*) are relatively common in the general population, with an estimated carrier frequency of approximately 2 % (about 1 in 50 individuals).
3. Variants associated with reduced blood ACE levels were estimated and measured to be carried by approximately 3.9 % of the general population, supporting the hypothesis that genetically determined ACE deficiency may represent a common and underrecognized biological contributor to late-onset AD susceptibility.
4. The publicly available *ACE* mutations–AD browser provides an integrated platform for exploring genotype–phenotype relationships and may support future research into ACE-dependent mechanisms of AD and related disorders.

## Data Availability

ACE mutations and AD database is freely available at https://ace-browser.com/.

## AVAILABILITY AND REQUIREMENTS

*ACE* mutations and AD database is freely available at https://ace-browser.com/.

## ETHICS

The protocols for the collection of human blood samples and subsequent whole-genome or whole-exome sequencing were reviewed and approved by the Ethics Committees of the Russian Gerontology Clinical Research Center (protocol no. 59, 13 September 2022) and the Ethics Committee of the Kulakov National Medical Research Center for Obstetrics, Gynecology and Perinatology (protocol no. 9, 22 October 2020). All corresponding procedures were carried out in accordance with institutional guidelines and the Code of Ethics of the World Medical Association (Declaration of Helsinki).

## ACKNOWLEDGMENTS

We thank those who have been a part of the referenced studies that form the basis of this paper and its accompanying online database.

## Supplementary Materials

**Table S1. Comprehensive catalogue of 1682 unique ACE coding variants.**

All variants identified from VarSome, dbSNP, and peer-reviewed literature, annotated with HGVS-compliant nomenclature, GRCh38 coordinates, rsIDs, gnomAD v4.1.0 allele frequencies, and consensus *in silico* pathogenicity scores (AlphaMissense, MetaRNN, EVE). Measured or estimated plasma ACE levels are included where available. Variants are stratified by functional category: signal peptide, missense, and frameshift/nonsense/indel.

**Table S2. Splice-site *ACE* variants and predicted splicing outcomes.**

All canonical and near-canonical splice-site variants identified within *ACE* (NM_000789.4) were analyzed using a tiered *in silico* framework combining SpliceAI, SAI-10k-calc, and SpliceVault RNA-seq data to predict the most likely splicing disruption mechanism for each variant. One variant (c.1709+5G>T) has been experimentally validated in patient-derived material; proposed mechanisms for the remaining variants are based on *in silico* predictions and transcriptomic evidence.

**Table S3. The 50 most frequent ACE coding variants by population allele frequency.**

The most common *ACE* variants are ranked by allele frequency derived from gnomAD v4.1.0, annotated with HGVS nomenclature, rsIDs, functional category, consensus *in silico* pathogenicity classification, and available plasma ACE phenotype data. This table provides a population-level summary of the most recurrent *ACE* alleles, including variants previously associated with altered enzymatic activity, shedding, or AD-related risk.

**Table S4. Amino acid residues and *ACE* variants predicted to influence catalytic activity.**

This table provides a curated list of residues in the N-domain (49 positions) and C-domain (29 positions) whose substitution is predicted or experimentally demonstrated to affect ACE enzymatic function, including zinc ion coordination, chloride binding, substrate accommodation, and inhibitor interactions. For each residue, all observed missense variants are reported alongside their *in silico* pathogenicity scores (AlphaMissense, MetaRNN, EVE, consensus), gnomAD v4.1.0 allele frequencies, available plasma ACE measurements, and structural or functional annotations derived from ACE–ligand complex structures, including complexes with amyloid-β42 fragments.

**Table S5. Amino acid residues and *ACE* variants predicted to influence dimerization.**

The table is divided into two parts: (a) 61 residues identified from structural analyses of X-ray and cryo-EM structures of full-length and truncated ACE dimers, including residues involved in glycan-mediated contacts at N82, inter-protomer packing at C-loop-2 and C-loop-3 regions, and D2–D4 subdomain interactions; and (b) 47 residues predicted to participate in dimerization based on docking simulations, including three C-domain residues. For each residue, all observed missense variants are reported with *in silico* pathogenicity scores (AlphaMissense, MetaRNN, EVE, consensus), gnomAD v4.1.0 allele frequencies, available plasma ACE measurements, and AD association where reported. Variants appearing in both parts reflect overlapping structural and docking-based predictions.

**Table S6. *ACE* variants associated with altered ectodomain shedding and circulating ACE levels.**

The table is divided into two parts: (I) 38 stop-codon variants in the mature ACE coding sequence that truncate the protein upstream of the transmembrane anchor, likely resulting in constitutive secretion of a soluble ACE ectodomain and elevated circulating ACE levels, with combined population frequency of approximately 0.05%; and (II) 41 missense variants located in the stalk and transmembrane anchor regions predicted to modulate proteolytic ectodomain release by the putative ACE secretase. For each variant, rsIDs, gnomAD v4.1.0 allele frequencies, in silico pathogenicity scores (AlphaMissense, MetaRNN, EVE, consensus), and available plasma ACE measurements are reported, with AD association indicated where previously described.

**Table S7. Plasma ACE activity measurements in carriers of *ACE* variants (RSMU cohort).**

The table lists 64 *ACE* variants for which blood ACE activity measurements were available from a total of 330 individuals, assessed by mAb 9B9-based precipitation followed by enzymatic assay with artificial substrates ZPHL and HHL. For each variant, GRCh38 genomic coordinates, HGVS protein nomenclature, amino acid position in the mature protein, rsID or literature reference, consensus pathogenicity classification, gnomAD v4.1.0 allele frequency, mean and median plasma ACE activity expressed as a percentage of healthy volunteer reference values (100%), and number of measured carriers are reported. The final row summarizes the cumulative allele frequency of variants associated with reduced plasma ACE activity.

**Table S8. *ACE* variants associated with reduced plasma ACE activity.**

The table lists 72 *ACE* variants consistently associated with decreased circulating ACE levels, organized into three sections: (I) 6 signal peptide variants; (II) 33 loss-of-function variants in the mature protein (frameshift, nonsense, and splice-site); and (III) 33 missense variants. For each variant, HGVS protein nomenclature, amino acid position in the mature protein, rsID or literature reference, AlphaMissense score (missense only), gnomAD v4.1.0 allele frequency, and median plasma ACE activity as a percentage of healthy volunteer reference values are reported, with AD association indicated where previously described. The combined allele frequency of all variants in this table is approximately 3.9% of the general population.

**Table S9. Monoclonal antibody-based conformational signatures for 19 *ACE* variants.**

The table lists 19 *ACE* variants for which mutation-specific conformational fingerprints were identified using a panel of more than 40 mAbs to epitopes in the N- and C-domains of ACE. For each variant, HGVS protein nomenclature, amino acid position in the mature protein, rsID or literature reference, *in silico* pathogenicity scores, gnomAD v4.1.0 allele frequency, median plasma ACE activity, and AD association are reported. The mutation marker columns specify the mAb pair or catalytic assay (ZPHL/HHL ratio) used to identify each variant, together with the mean binding ratio expressed as percentage of control. For variants causing gross conformational changes (Y465D, R532W) or domain-selective effects (Q1069R), multiple antibodies were required for characterization.

